# Predicting pneumothorax after lung biopsy from pre-operative imaging using a deep convolutional neural network

**DOI:** 10.1101/2022.03.07.22271554

**Authors:** Seong Jun Jang, Bradley B. Pua, George Shih

## Abstract

**Background:** Pneumothorax remains one of the most common complications after computed tomography (CT)–guided lung biopsies. Radiographic features including bullae and nodule size are possible markers for post-biopsy pneumothorax. We determine whether a convolutional neural network (CNN) can accurately predict a pneumothorax after lung biopsy based on pre-operative imaging alone.

**Methods:** With institutional review board approval, we retrospectively evaluated 3,822 patients who underwent a CT-guided lung biopsy between 2011 to 2019. Two image sets were created with CT scout images (1300 patients, 650 pneumothoraces) and chest x-rays (CXR) taken within three months pre-procedure (884 patients, 140 pneumothoraces). Using pre-operative images, CNNs of varying layer depths were trained using transfer learning to predict the development of a pneumothorax post-biopsy. Performance against models were compared using sensitivity analysis and the McNemar’s test.

**Results:** The CNN models trained with CT scout images performed near chance. However, the models performed better with CXR radiographs taken within three months pre-biopsy. For the anterior-posterior view, sensitivity was 0.40, specificity was 0.89, PPV was 0.43, and NPV was 0.87 (AUC = 0.67). For the lateral view, sensitivity was 0.40, specificity was 0.80, PPV was 0.32, and NPV was 0.86 (AUC = 0.65). Increasing CNN layers did not affect performance (p > 0.05).

**Conclusion:** Chest radiographs taken within three months of lung biopsy may provide important radiographic information for CNNs to assess pneumothorax risk in patients prior to CT-guided lung biopsies. However, more baseline and standardized CXRs before biopsies are necessary to create a robust model for clinical application.

## Introduction

Computed tomography (CT)-guided percutaneous lung biopsies serve an important role in the diagnosis and management of pulmonary nodules^1^. Although generally regarded as a safe and reliable procedure, CT-guided lung biopsy can result in multiple complications including pneumothorax, pulmonary hemorrhage, air embolism, and tumor seeding^2,3^. Notably, pneumothorax is one of the most common complications, with a recent systematic review reporting a pneumothorax rate of 25.9% and a chest tube insertion rate of 6.9% after lung biopsy in over 20,000 patients^4^.

Given the high complication rate of pneumothorax and chest tube placement after lung biopsies, investigations have aimed to identify pertinent risk factors to detect patients at higher risk and to subsequently minimize the chance of complications. The rate of pneumothorax after lung biopsy is affected by a variety of factors that are dependent on the characteristics of the lung, nodule, needle, and procedural approach^2–5^. Likewise, the presence of severe obstructive pulmonary disease is associated with an increased incidence of pneumothorax requiring chest tube drainage post-biopsy ^4–6^. Additionally, studies have shown that the radiographic characteristics of the lung such the presence of bullae^7^ and obstructive lung disease^8^ as well as lesion characteristics including smaller, deeper lesions have a greater risk of pneumothorax^7,9^. The presence of obstructive airway disease on radiographs is also associated with a significantly higher complication rate of pneumothorax (42%) versus no sign of obstructive airway disease (19%), emphasizing the importance of using radiograph features as possible markers for risk stratification in individuals undergoing lung biopsies^8^. While many studies have been published on possible risk factors associated with the development of pneumothorax, they are often conflicting or inconclusive in isolating specific risks.

With the multitude of factors associated with increased pneumothorax risk after lung biopsies, machine learning approaches have been recently utilized to better predict complication outcomes. In particular, a study determined which features of patient data were strongly associated with the incidence of a pneumothorax after lung biopsies, finding that patient and procedure characteristics such as age and needle size were important factors in pneumothorax incidence^10^. Another deep learning model based on clinical and demographic data predicted a pneumothorax after lung biopsy with 90% sensitivity and 81.6% specificity^11^. As these studies demonstrate, it is possible to apply machine learning algorithms to predict complications after lung biopsies.

Baseline radiographic images of patients provide another important set of data that has yet to be explored for post-biopsy pneumothorax prediction with machine learning methods. Regarding the role that radiographic images play for pulmonary nodules, chest radiographs or lung CT screening often detect the presence of a pulmonary nodule prior to a biopsy. Nodule or clinical characteristics then further warrant a CT-guided lung biopsy for pathological evaluation. With any CT scan, scout images are taken prior to the CT-guided lung biopsy to plan slices, and scout views can be used to identify significant radiographic findings in patients^12^. Thus, scout views, in addition to chest radiographs, offer two sources of pre-operative radiographic data for patients undergoing a biopsy.

In this study, we aim to utilize a machine learning approach to identify patients at risk for pneumothorax after lung biopsies based on pre-operative radiographic data. We hypothesize that a trained deep convolutional neural network (CNN) will be able to accurately predict the complication of a pneumothorax after lung biopsy. We will use a pre-weighted CNN for an image-based machine learning approach. This process, where a pre-weighted neural network optimized for a generalized task such as image classification is developed for another classification scheme, is referred to as transfer learning. It has been applied in other medical fields such as ophthalmology for the detection of diabetic retinopathy^13^ and dermatology for the classification of malignant vs non-malignant nevi with high accuracy and performance against that of trained physicians^14^. Furthermore, studies have also applied CNNs in radiology for the detection of abnormal CXRs that warrant clinical investigation^15^. Like these previous works, our goal is to also use a CNN approach to predict an outcome based on image data as it specifically relates to CT-guided lung biopsies performed by interventional radiologists.

## Methods

### Study Cohorts

Institutional review board approval was acquired under a protocol designated to assess the accuracy and safety of image-guided lung biopsies. All patients who underwent a lung biopsy from 2011 to March 2019 were originally identified (3,822 patients) from the Weill Cornell Lung Cancer Screening Program. In this group, 17.8% of patients had a pneumothorax (682 patients). Outcomes of no pneumothorax versus pneumothorax were all established by interventional radiologists based on imaging and the post-operative course of patients.

Of the identified patients, two separate study cohorts were made for modeling with pre-biopsy chest x-rays (CXR) or computed tomography (CT) scout images. Inclusion criteria for the CXR cohort included all patients who received a percutaneous lung biopsy and had a pre-operative chest radiograph at most three months prior to the procedure (884 patients). Three months was chosen as a conservative time frame to best capture patient lung status on radiographs at a date close to the biopsy. In this group, 140 patients (15.8%) had a pneumothorax. Inclusion criteria for the CT scout image cohort included all patients who received a percutaneous lung biopsy and had scout views available on their CT-guided lung biopsy. For this cohort, the first 650 patients (50%) who developed a pneumothorax were selected and matched by sex to 650 other patients who did not develop a pneumothorax to create a final cohort of 1300 patients. Images were also separated by view for CT scout images (anterior-posterior and lateral) as well as for chest radiographs (anterior-posterior and lateral), creating a total of four unique datasets for training. Every image was anonymized, and the authors were blinded during model training with access to only anonymized imaging data.

### CT-Guided Lung Biopsy

The lung biopsy protocol was standardized across for the duration of the entire study. All biopsies were performed by either a fellowship-trained interventional radiologist faculty, an interventional radiology resident or fellow under the guidance of fellowship-trained faculty, or by a physician assistant with more than 15 years of experience under the guidance of faculty. A scout image and repeat low dose CT surrounding the abnormality was performed at the time of each biopsy. Based on the CT scan, a mark was placed on the patient skin for the area of entry. The trajectory was chosen to limit the amount of lung traversed, to limit the number of fissures crossed, and to avoid vital structures and areas of high emphysema. The procedure was conducted with the patient sedated or with local anesthesia. Patients underwent general anesthesia secondary to medical comorbidity. The needle was passed under CT guidance and the number of passes and samples were determined in conjunction with an on-site cytopathologist to increase yield and diagnostic accuracy of the lung biopsy. After the needle was removed, a limited CT was performed to assess for immediate complications. After biopsy, the patient was taken to the recovery room, and placed in a position with the side of biopsy facing the ground for two hours. Chest x-ray was performed at zero and two hours after completion of biopsy. The patient was discharged home if there were no indication of complications on radiographs that required the escalation of care.

### Deep Learning Algorithm and Training

The fastai (V2.3.0) deep learning library^16^ was utilized for training the CNNs, and ResNet models with pre-trained weights for image analysis were chosen as the architectures for both imaging modalities. The loss function, hyperparameters, and learning rate were all determined using fastai’s model optimization functions. Batch sizes were chosen as 16 for ResNet-50 and 32 for ResNet-34 and ResNet-18. For training, the datasets were randomly split into training and validation sets in an 8:2 ratio. Data augmentation was applied to the training set with rotation (max 10 degrees), zoom (max 1.2), and wrap transforms with the fastai library. Images were also normalized based on DICOM pixel data. Furthermore, given the relatively small size of the CXR datasets, we employed oversampling of the pneumothorax cohort to ensure that training data had a 50:50 split of pneumothorax and no pneumothorax outcome images. In this process, we ensured that no patient image or images from the same patient appeared in both the training and validation set as having the data in both sets could train the model to falsely learn to identify individual patients or pictures due to data leakage. Finally, validation sets were predetermined to have a pneumothorax rate of 20% for the CXR dataset to reflect the incidence reported in literature.

The training and validation loss were tracked through various epochs during model training to ensure CNN learning. The best models were chosen upon the point at which the validation loss began to increase to avoid overfitting the data to the training set. The model was saved at that epoch and used for analysis. Several ResNet architectures (−18, -34, -50) were also trained to compare results based upon adding more layers for the same validation set. The models were trained on NVIDIA CUDA with 12 gigabytes of RAM and on PyTorch.

### Algorithm Testing and Statistical Analysis

After training, model performance was tested on the validation sets. The number of true positives, false positives, true negatives, and false negatives were analyzed, and the sensitivity, specificity, positive predictive value, and negative predictive value of the algorithm were calculated. For each image modality and view, the three ResNet architectures were statistically compared against each other using the McNemar test with the same validation set. The area under the receiver operating characteristic (ROC) curve was also determined to demonstrate performance. The model with the best performance was selected as the final model for pneumothorax outcome prediction after lung-biopsy based on CXR or CT scout images. Class activation maps were also created to qualitatively analyze possible features utilized by the CNN to generate predictions.

## Results

### Patient Characteristics

Patient demographics are outlined for each patient cohort in Table 1. In total, there were 1300 patients in both CT image groups. In the CXR anterior-posterior (AP) view cohort, there was a total of 679 patients. In the CXR lateral view cohort, there was a total of 392 patients given that a lateral view of the CXR was not indicated for many patients at time of imaging. There were no significant differences between sex and weight in all cohorts in this study.

**Table 1:**
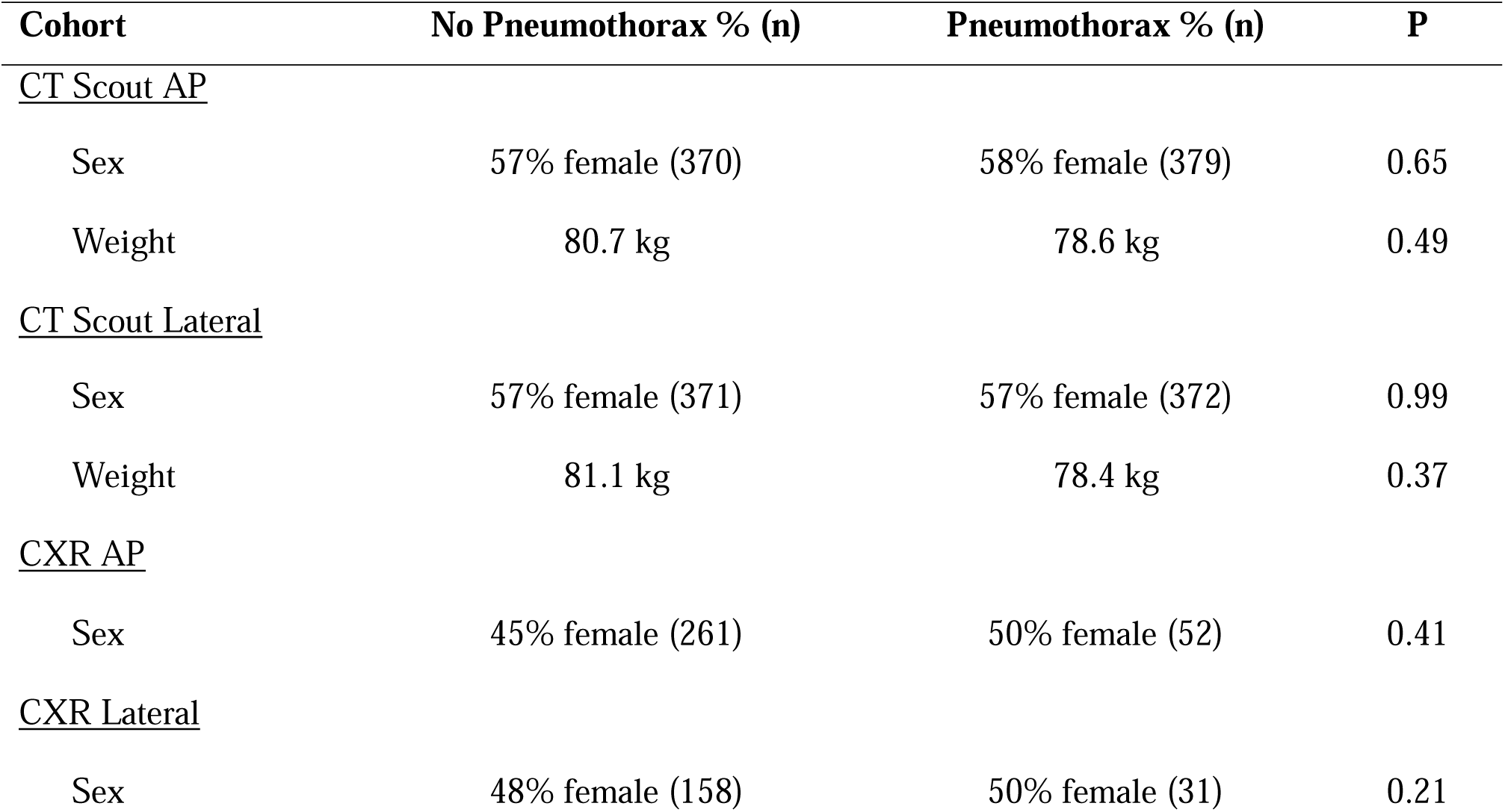
**Demographic characteristic of all cohorts based upon outcome of pneumothorax and no pneumothorax after lung biopsy**

### CT

For AP view scout CT images, the area under the curve (AUC) of the receiver operating characteristic (ROC) curve was 0.61 for ResNet-18. The AUC was 0.59 for ResNet-34 and 0.56 for Resnet-50 (Figure 1a). However, the performance of the ResNet-18 model was not significantly better than that of ResNet-34 (p = 0.68) or ResNet-50 (p = 0.75). The confusion matrix using ResNet-18 is depicted in Figure 2a with 0 representing outcome of no pneumothorax and 1 representing outcome of pneumothorax. From the validation set, the model had a sensitivity of 0.40, specificity of 0.73, positive predictive value of 0.61, and a negative predictive value of 0.54, leading to an accuracy 0.57 with the given pneumothorax prevalence of 0.5 (Table 2).

**Table 2:** Performance metrics of all models for each imaging modality and view.

| **Model** | **<u>Sensitivity</u>** | **<u>Specificity</u>** | **<u>PPV</u>** | **<u>NPV</u>** | **<u>AUC</u>** |
| --- | --- | --- | --- | --- | --- |
| **<u>CT AP</u>** |  |  |  |  |  |
| Resnet-18 | 0.40 | 0.73 | 0.61 | 0.54 | 0.61 |
| Resnet-34 | 0.44 | 0.65 | 0.57 | 0.53 | 0.59 |
| Resnet-50 | 0.36 | 0.74 | 0.54 | 0.53 | 0.56 |
| **<u>CT Lateral</u>** |  |  |  |  |  |
| Resnet-18 | 0.46 | 0.52 | 0.48 | 0.46 | 0.52* |
| Resnet-34 | 0.47 | 0.54 | 0.52 | 0.49 | 0.55 |
| Resnet-50 | 0.40 | 0.72 | 0.60 | 0.53 | 0.55* |
| **<u>CXR AP</u>** |  |  |  |  |  |
| Resnet-18 | 0.43 | 0.77 | 0.29 | 0.86 | 0.65 |
| Resnet-34 | 0.39 | 0.89 | 0.43 | 0.87 | 0.67 |
| Resnet-50 | 0.30 | 0.88 | 0.35 | 0.85 | 0.67 |
| **<u>CXR Lateral</u>** |  |  |  |  |  |
| Resnet-18 | 0.27 | 0.83 | 0.27 | 0.83 | 0.63 |
| Resnet-34 | 0.40 | 0.80 | 0.32 | 0.85 | 0.66 |
| Resnet-50 | 0.53 | 0.74 | 0.32 | 0.88 | 0.65 |

\* ResNet-50 performed significantly better than ResNet-34 on the CT Lateral Images

**Figure 1:**
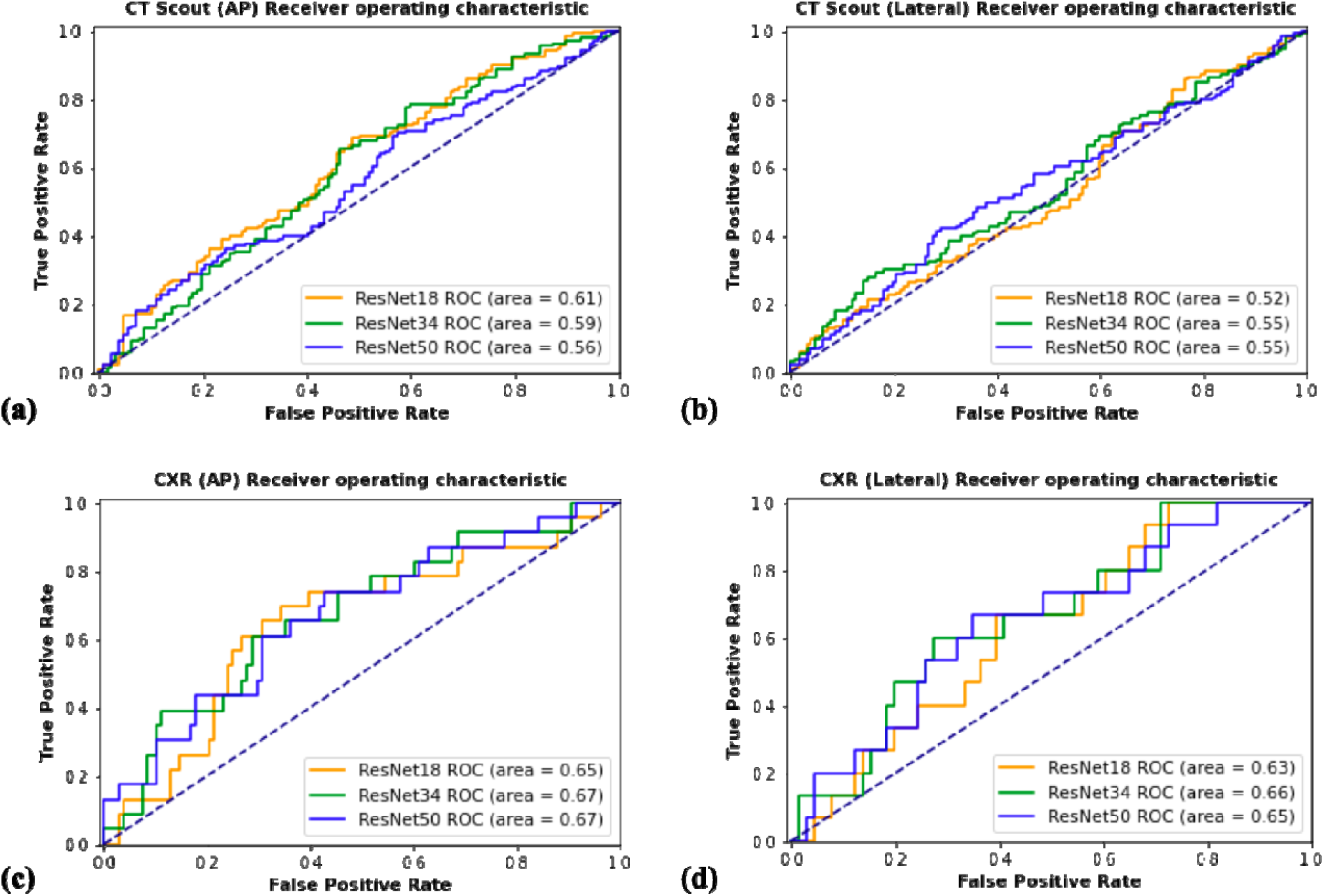
AUC-ROC results with all models. A) CT scout radiographs in the anterior-posterior (AP) view. No significant differences between the model performances. B) CT scout radiographs in the lateral view. ResNet-50 performed significantly better than ResNet-18 on the same validation set. C) CXR radiographs in the anterior-posterior (AP) view. No significant differences between the model performances. D) CXR radiographs in lateral (AP) view. No significant differences between the model performances.

**Figure 2:**
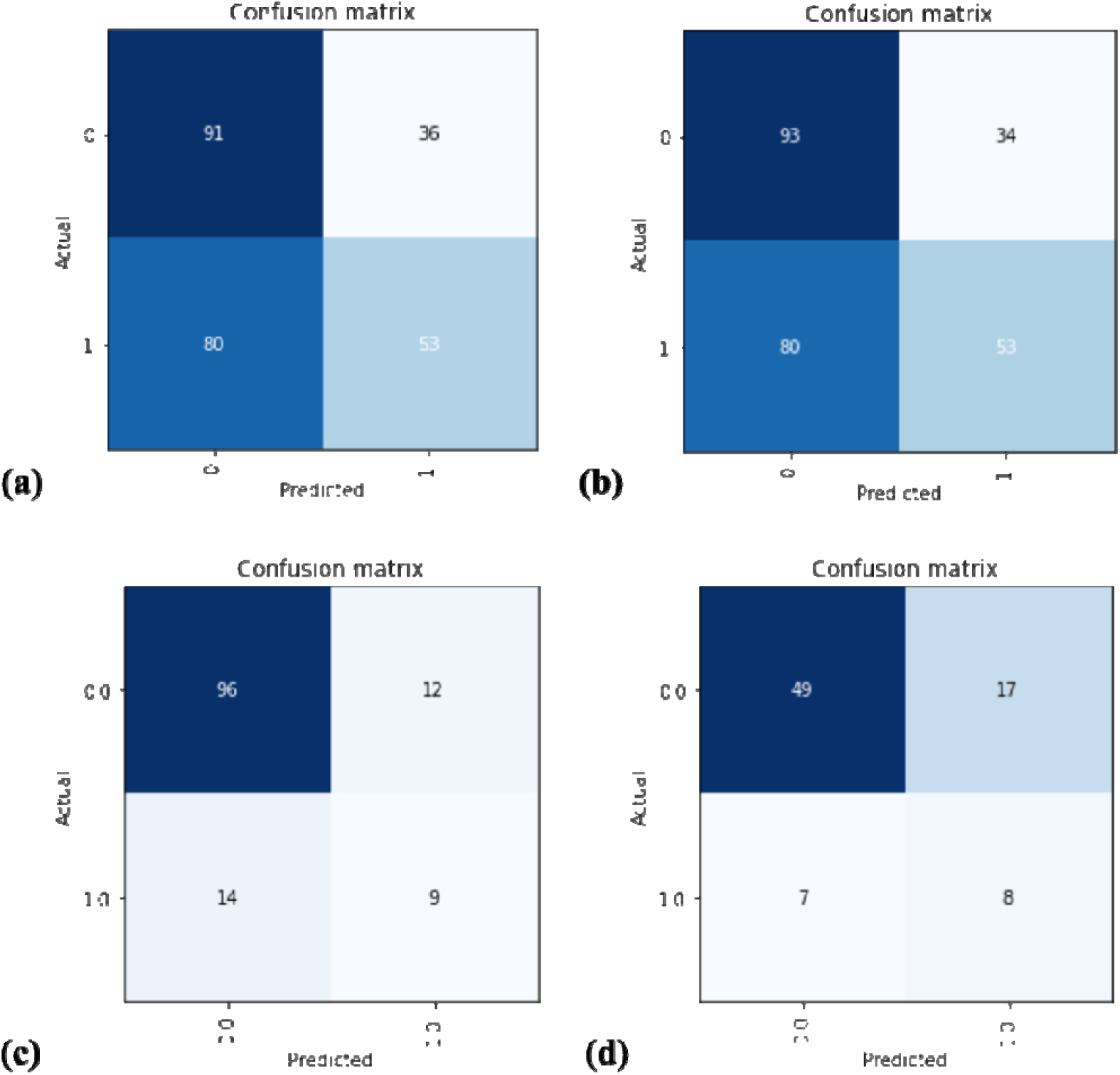
Confusion matrices of all models. A) CT scout radiographs in the anterior-posterior (AP) view with ResNet-18. B) CT scout radiographs in the lateral view with ResNet-50. C) CXR radiographs in the anterior-posterior (AP) view with ResNet-34. D) CXR radiographs in lateral (AP) view with ResNet-50. The CXR AP view model had higher specificity (0.89) but lower sensitivity (0.39). Both CXR AP and lateral models had high negative predictive values (AP 0.87, lateral 0.88).

For lateral view scout CT images, the AUC of the ROC was 0.52, 0.55, and 0.55 for ResNet-18, -34, and -50, respectively (Figure 1b). ResNet-50 performed significantly better compared to ResNet-18 on the same validation set (p = 0.038) however, it did not perform significantly better than ResNet-34 and there was no significant difference between the performance of ResNet-18 and ResNet-34 (Table 2). For ResNet-50, which had the highest AUC, the sensitivity was 0.40, specificity was 0.72, positive predictive value was 0.60, negative predictive value was 0.53, with an accuracy of 0.56 on the validation set (Figure 2b).

### CXR

For CXR images in the AP view, the AUC of the ROC curve was 0.65, 0.67, and 0.67 for ResNet-18, -34, and -50, respectively (Figure 1c). ResNet-50 and ResNet-34 both had an accuracy of 76% on the validation set. However, one did not perform significantly better compared to the another (p = 0.68) (Table 2). Furthermore, ResNet-50 and ResNet-34 did not perform better than ResNet-18 (p = 0.07, and p = 0.15), which had an accuracy of 0.70 with the same validation set (Table 2). For ResNet-34, which had the highest AUC, the sensitivity was 0.40, specificity was 0.89, PPV was 0.43, and NPV was 0.87 on the validation set with a pneumothorax prevalence of 20% (Figure 2d).

For CXR images in the lateral view, the AUC of the ROC curve was 0.63, 0.66, and 0.65, for ResNet-18, -34, and -50, respectively. ResNet-34 did not perform better than ResNet-18 (p = 1.00) or ResNet-50 (p = 0.84) (Table 2). For ResNet-34, the sensitivity was 0.4, specificity was 0.80, PPV was 0.32, NPV was 0.86, and accuracy was 0.70 on the validation set with a pneumothorax prevalence of 20% (Figure 2d).

## Discussion

In this work, we investigated the use of a convolutional neural network for the identification of pneumothorax outcome after CT-guided lung biopsies based on pre-operative imaging. We utilized both scout CT images taken immediately before biopsies and chest radiographs taken within 3 months before biopsy. Our CNN model performed best with preoperative chest radiographs in the AP view (AUC = 0.67) with a sensitivity of 0.40 and specificity of 0.89, as well as a PPV of 0.43 and an NPV of 0.87 on the validation set.

Importantly, this is the first model to our knowledge to predict post-biopsy pneumothorax outcomes using pre-biopsy imaging alone. Previous works have focused on the identification of pneumothorax post-biopsy on chest radiographs. These studies have demonstrated that models can achieve sensitivities of 61.1% to 70.5%, specificities of 93.0% to 97.0%, and an AUC of 0.90 to 0.94 in detecting pneumothorax on chest radiographs post-operatively^17,18^. These findings are in line with our model, which exhibited a lower sensitivity over specificity. Although our model did have lower performance metrics compared to these previous models, this is expected given that previous models were detecting the presence of an already-defined visual finding on a given radiograph (i.e. visible pleural edge, radiolucent peripheral space). Critically, our model’s task was to predict the outcome of a pneumothorax before it occurred based on pre-operative imaging. Rather than identifying pneumothoraces, our model’s task was to discern between radiographs that have increased risk of developing pneumothoraces before a lung biopsy based on features that it would learn on its own. Class activation maps were created for the CNN model with AP chest radiographs to help discern features which the model may use in its predictions (Supplemental 1 and 2). They suggest that the visible opacities or nodules on the radiographs were features utilized in the final prediction for pneumothorax. However, more discrete analysis with the activation maps is necessary to make conclusions regarding radiographic features associated with a pneumothorax post-biopsy.

The best CXR models performed better at predicting an outcome of no pneumothorax compared to predicting an outcome of pneumothorax (high specificity, lower sensitivity). This can be explained by a multitude of factors. Namely, the prevalence of pneumothorax in our training set was 15.8%, which was lower than that reported from previous studies^4,17,18^. As such, the model may not have had enough samples of radiographs with a pneumothorax outcome to accurately discern features to base its predictions on. Indeed, training the model with this imbalanced training set originally led to the model to predict an outcome of no pneumothorax on all images, achieving falsely elevated accuracies close to ∼84% (100%-15.8%) with zero sensitivity. To curtail this imbalance in data, we augmented and oversampled the images with a pneumothorax outcome to create an even 1:1 ratio of no pneumothorax and pneumothorax in the training set. Doing so led to the current sensitivities of 0.39 and 0.53 in the best CXR models, compared to zero. This improvement suggests that higher sensitivity could be achieved with more cases of pneumothorax as it is likely that model did not have enough samples to generalize features on. Likewise, higher specificity could also be achieved with more cases of no pneumothorax as well given the relatively small size of this dataset.

We also demonstrate that models trained with chest radiographs exhibit superior performance compared to CT scout images in predicting pneumothoraces pre-lung biopsy. CT scout images are traditionally used to aid with patient and field positioning before CT scans. Although previous studies have advocated for its use in routine inspection for pathologic detection^12^, these images are of lower-resolution and may not provide enough information to the deep neural network for the task at hand. In contrast, chest radiographs are of higher resolution and can provide better data for the convolutional neural network to identify features that one might find in patients at risk for pneumothorax such as emphysema or scarring. Unfortunately, chest radiographs are currently not the standard-of-care in patients planned to undergo lung biopsies. Rather, the chest radiographs selected in this study were taken for other indications such as fever and chest pain. Nonetheless, despite the varying array of clinical indications, we find that a chest x-ray within 3 months of a lung biopsy can provide useful data for a convolutional neural network to discern a post-operative outcome, pre-operatively, compared to a scout CT image.

With more data to increase the sensitivity and specificity of this deep learning model, there are many potential clinical applications of this work. Although our work focused on pneumothorax, other outcomes after lung biopsies including pulmonary hemorrhage and hemothorax^2^ could also be investigated. If a CNN can predict other complications using pre-operative chest radiographs, then perhaps a chest radiograph provides enough information to a CNN to warrant routine application in assessing operative risk in CT-guided lung biopsies. This would aid operators in identifying patients at risk and in planning procedures with more scrutiny to avoid such complications. However, more data is first necessary to create a robust model and prospective studies are necessary to determine the effectiveness of applying this CNN in daily practice.

The results of this model can also be augmented with the works of other machine learning approaches in CT-guided lung biopsies. A recent study utilized a neural network approach with patient demographic and clinical information to predict complications after CT-guided lung biopsies^19^. These models can be combined with our model in an ensemble network to better predict lung biopsy complications using demographic, clinical and now radiographic data. Furthermore, this current study separated the datasets based on image modality (CT vs CXR) and image view (AP vs Lateral). Future investigations will involve creating ensemble models where the neural network creates predictions for each type of image and view for one patient and subsequently makes a combined, weighted prediction for a final decision of pneumothorax or no pneumothorax. Finally, the results of our study can be statistically compared against the results of trained radiologists and interventional radiologists to predict pneumothorax after lung biopsies based on pre-operative chest radiographs. Doing so would validate the increased effectiveness of this model in stratifying patients based on risk through imaging alone.

Various ResNet depths were also explored in this study. Recently, the lack of external, multi-institutional, prospective validation sets has raised the concern of non-generalizability and model overfitting to an institution’s data in multiple studies^20^. In this current study, architectures with lower number of layers were utilized to reduce the chance of overfitting the model to our data given the relatively small sample size. Despite using lower numbers of layers to avoid overfitting and promote generalization, the model did not suffer from the loss of its ability to extract features to predict pneumothorax outcomes. This result is in line with studies that show networks with lower complexity and number of layers produce comparable, and often more efficient, results in medical imaging task^21^.

There were several limitations. Firstly, we utilized a relatively small sample size to train the models. Despite this limitation, we were able to create a neural network model with relatively high specificity and NPV based on pre-operative AP chest radiographs. Secondly, only CXR AP and lateral views of the chest were used. PA views were available in a limited number of patients and thus not included. Thirdly, we created the validation set to better match the pneumothorax prevalence reported in previous studies (20%) for the CXR models despite having a smaller cohort size for deep learning. Finally, all the data in this study was confined to one institution and future studies will be necessary to validate this model with prospective data from different facilities for external validation.

## Conclusion

A deep convolutional neural network achieved higher performance in predicting post-biopsy pneumothorax using pre-operative chest radiographs taken within three months of biopsy compared to CT scout images taken directly before biopsy. More radiographic training data and external validation of this model is necessary for its application in clinical workflow. However, future models have the potential to stratify patients based on pneumothorax risk before biopsy and lower the clinical threshold for increased planning and evaluation to decrease the risk of post-biopsy pneumothorax.

## Data Availability

All data produced in the present work are contained in the manuscript.

## Acknowledgements

We thank Meghan Cahill, Deidre Sullivan, and Benjamin Cobb for aiding in the acquisition of patient imaging utilized in this work. We thank Kurt Teichman for his technical support throughout the project. Finally, we thank the Weill Cornell Medical College Area of Concentration program for providing guidance on this work and the Department of Interventional Radiology as well as the Department of Radiology at the New York Presbyterian Hospital for sponsoring this project.

## Supplemental Figures

**Supplementary Figure 1:**
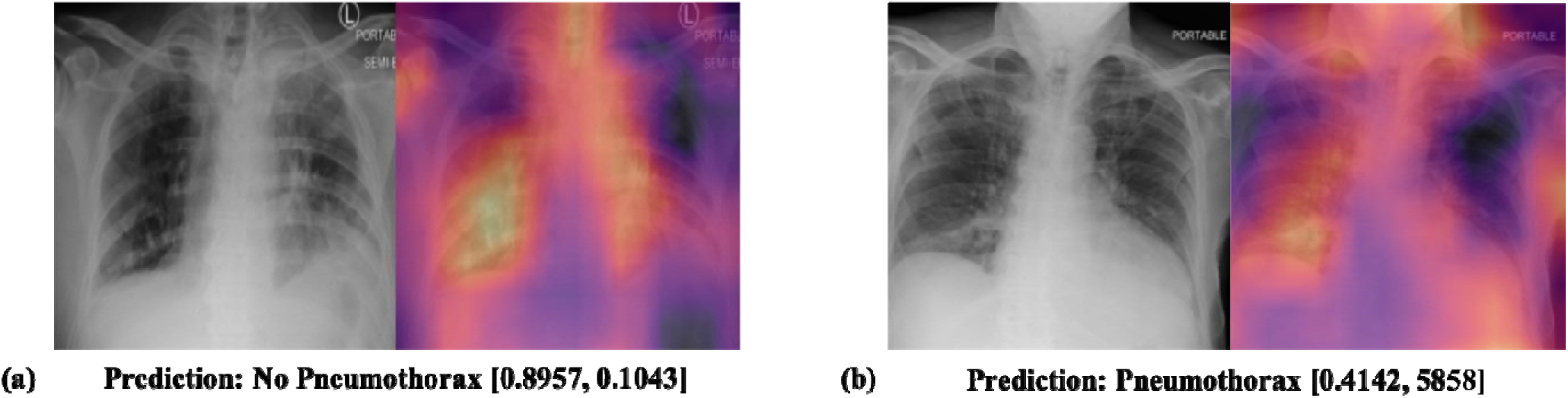
Class activation maps of model created with ResNet-34 on AP CXRs with ground truth of no pneumothorax. Final predictions with softmax represented in brackets [no pneumothorax, pneumothorax]. A) Example of correctly predicted image in the validation set highlighting right lower lung. B) Example of incorrectly predicted image in the validations set highlighting abnormality in right lower lung

**Supplementary Figure 2:**
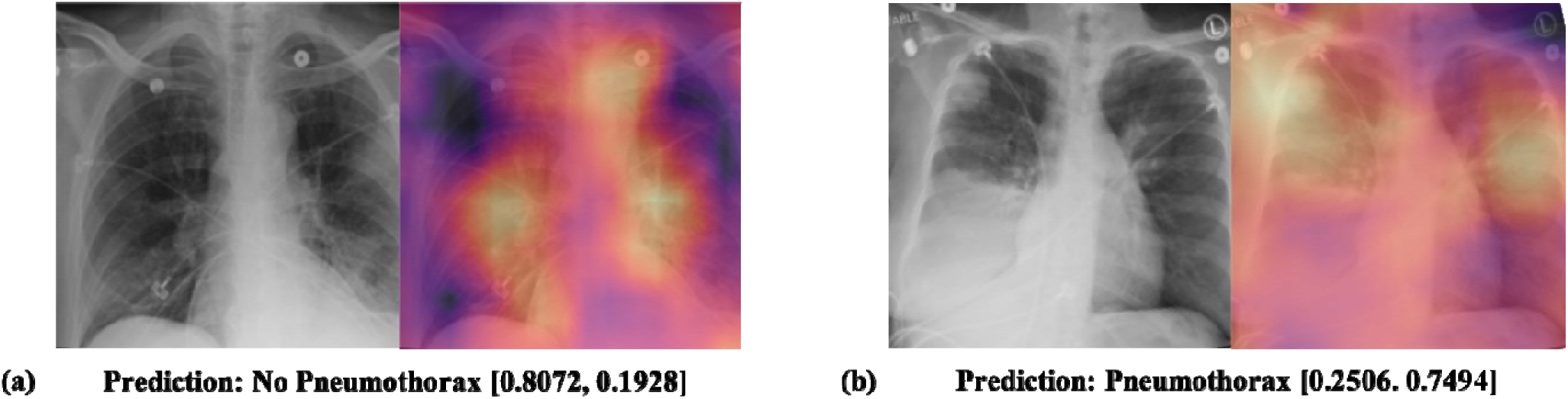
Class activation maps of model created with ResNet-34 on AP CXRs with ground truth of pneumothorax. Final predictions with softmax represented in brackets [no pneumothorax, pneumothorax]. A) Example of incorrectly predicted image in the validation set highlighting areas in bilateral lungs. B) Example of correctly predicted image in the validations set highlighting possible features in the bilateral lungs

**Supplementary Figure 3:**
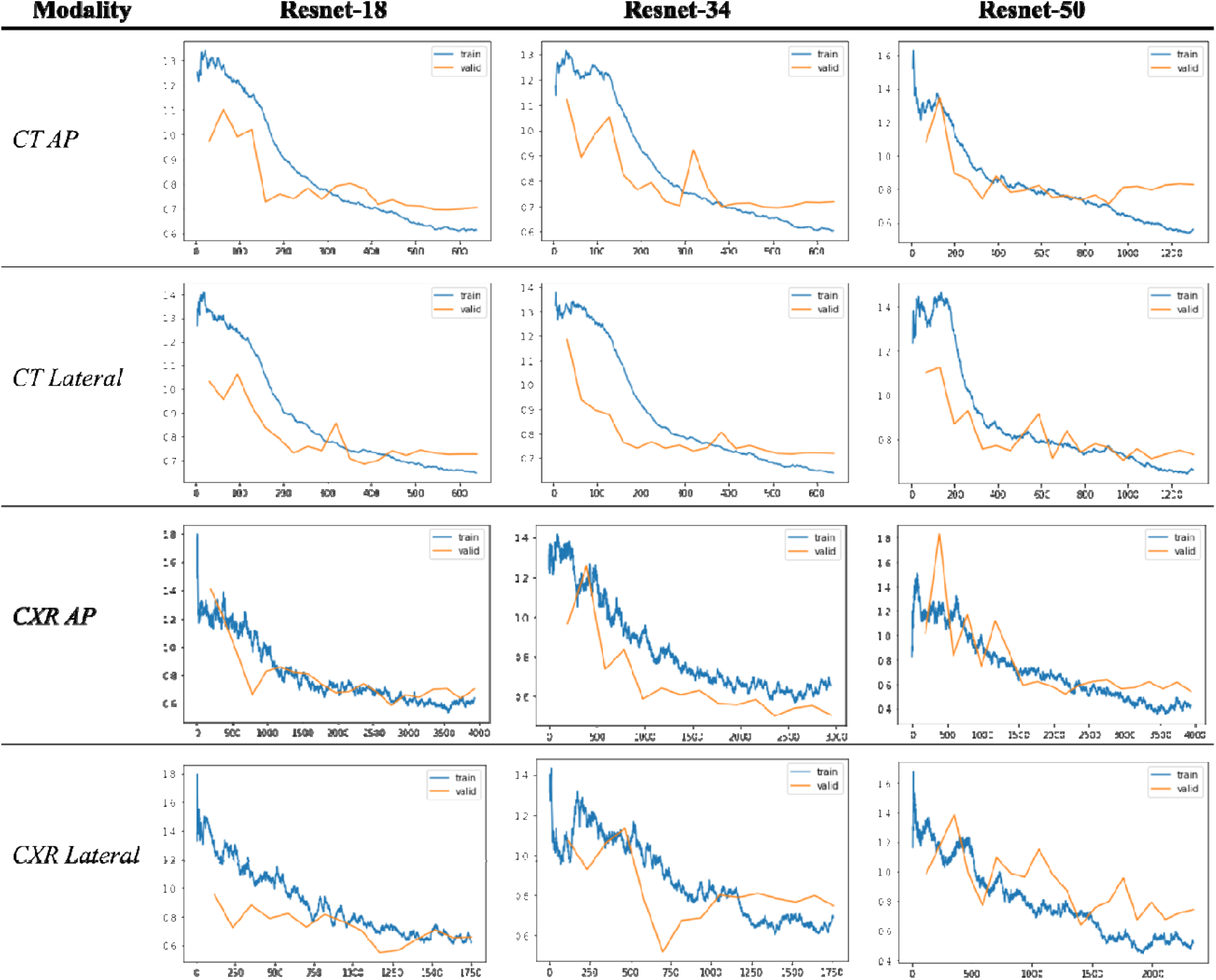
Training and validation loss over model training. X axis represents the number of images that the model is learning from over time. Y axis represents the cross-entropy loss over time for both training and validation sets. Gradual increase in loss over time may represent overfitting to the training set and final models are chosen based upon the epoch with the lowest loss before this increase.

